# Antibody response after a single dose of ChAdOx1-nCOV (Covishield™®) vaccine in subjects with prior SARS-CoV2 infection: Is a single dose sufficient?

**DOI:** 10.1101/2021.06.15.21258346

**Authors:** Biswajyoti Borkakoty, Mandakini Das Sarmah, Chandra Kanta Bhattacharjee, Nargis Bali, Gayatri Gogoi

## Abstract

It is crucial to know whether a single dose of vaccine against SARS-CoV-2 is sufficient to elicit immune response in previously infected people in India. A total of 121 participants (baseline seropositive 46 and seronegative 75) were included to study the immune response to ChAdOx1-nCOV (Covishield) vaccine in previously infected or uninfected people. IgG antibodies were estimated at three different time intervals, i.e. pre-vaccination, 25-35 days post 1st vaccination and 25-35 days post 2nd vaccination. The IgG antibody titre was significantly high among previous seropositive subjects with single dose of vaccine compared to seronegative group with both doses of vaccine respectively (4.59±1.04 vs 3.08±1.22, p-value: <0.0001).

In conclusion, a single dose of Covishield™® vaccine might be sufficient to induce an effective immune responsein subjects with prior SARS-CoV2 infection. Stratifying vacinees based on their SARS-CoV2 IgG antibody titre before vaccination would help in meeting the increasing vaccine demand and could be effective to circumvent further wave of the pandemic in India.

## Introduction

Only 3.2 percent of India’s 1.4 billion population have been immunized against the coronavirus disease 2019 (COVID-19) as of 2^nd^ June, 2021 [1]. The number of reported COVID19 cases in India till date is 28, 441, 986 with 338,013 confirmed deaths [1]. Two vaccines namely Covishield (ChAdOx1-nCOV), and Covaxin(BBV152 whole virion inactivated vaccine) have been approved under emergency use authorization in India. The immunization programme for COVID-19 started on January 16 2021, with healthcare workers followed by frontline workers, and elderly persons being the first beneficiaries of the campaign. Mass vaccination for people above 18 years of age has also been initiated. As of 2nd June 2021, in India 43.6 million people have been fully vaccinated, and 213 million have received the first dose of the currently approved vaccines [1].

In a country like India, which is currently experiencing the second wave of the COVID-19 pandemic, with thousands of cases reported daily, mass immunization measures are urgently needed. The country’s present vaccine manufacturing capacity was unable to fulfil the increasing demand, resulting in a vaccine shortage. Prioritizing vaccinees and categorically vaccinating the non-immune population is the need of the hour. In people with previous SARS-CoV-2 exposure, vaccine studies reported worldwide demonstrated significant antibody titers post first dose of Pfizer or Moderna RNA vaccine [2-5]. Despite the fact that the trials only covered a small number of people, the main outcomes are similar: people with pre-existing immunity generated uniformly high antibody titers. In the light of the studies on other vaccines, the present study was conducted from January 2021 to April 2021, to explore the dynamics of the immune response to the Covishield (ChAdOx1-nCOV) vaccine (the major vaccine being used in India), in subjects who had previously been infected with SARS-CoV2 (baseline seropositive), compared to naive subjects (baseline seronegative).

## Materials and Method

A total of 121 participants with or without pre-existing SARS-CoV-2 immunity (baseline sample seropositive 46 and seronegative 75) were included in the present study. Before enrollment, signed consent was obtained from all the participants. The serum IgG antibodies against SARS-CoV2 were detected using a commercial ELISA kit approved by the FDA for emergency use authorization (SCoV-2 Detect IgG ELISA, InBios, USA, Cat No: COVE-G). Serum IgG antibodies were estimated at three different time intervals; just before vaccination (baseline sample) followed by 25-35 days post 1st vaccine dose (before receiving the 2nd dose) and again 25-35 days after 2nd vaccine dose.

## Ethical approval

The Indian Council of Medical Research- Regional Medical Research Centre North Eastern Region, Dibrugarh’s institutional ethics committee approved the study.

## Results and discussion

The average age of the 121 enrolled subjects was 33.7 years ± 11.9; age range: 18-75 years, males 45.4% and female 54.6%. The optical density (OD) measured at 450 nm in baseline seropositive post first dose (4.59±1.04 vs 2.98±1.53 p-value <0.0001) and post-second dose (4.31±0.89 vs 3.08±1.22, p-value <0.0001) was significantly higher compared to the baseline seronegative group (Figure 1A). Further, among the seropositive cases, the 2nd dose of vaccine did not raise the antibody titre any higher compared to the 1st dose (4.59±1.04 vs 4.31±0.89, p-value 0.17). Interestingly it was observed thatthe IgG antibody titre was considerably higher in previously seropositive participants who received only one dose of vaccine compared to the seronegative group who received both doses of vaccine (4.59±1.04 vs 3.08±1.22, p-value <0.0001) (Figure 1B). This study suggests that a single dosage of the vaccine may be enough to produce an efficient immune response in previously infected patients, even those with moderate illness. Despite the fact that the study did not look at cell-mediated immunity, it does provide useful information on antibody immune responses in previously infected and uninfected COVID-19 participants after vaccination. Given the limited availability of the second dose in a densely populated country like India, this is of utmost importance. Our findings are consistent with recent findings on the Moderna mRNA/Pfizer/BioNTech COVID-19 vaccines, which reported that vacinees who had previously been infected had considerably higher antibody titers than naive recipients [2-5].To the best of our knowledge, there has been no published report on the Covishield vaccine’s immune response to a single dose in previously exposed COVID-19 individuals. Despite the fact that our study only included a small number of participants, it gives crucial preliminary data on the dynamics of the antibody response to the Covishield vaccination, which is now the most widely used vaccination in India. The persistence of SARS-CoV-2immunological memory is confirmed by the quick and robust immune response to vaccination in prior SARS-CoV2 infected subjects.

**Figure 1.**
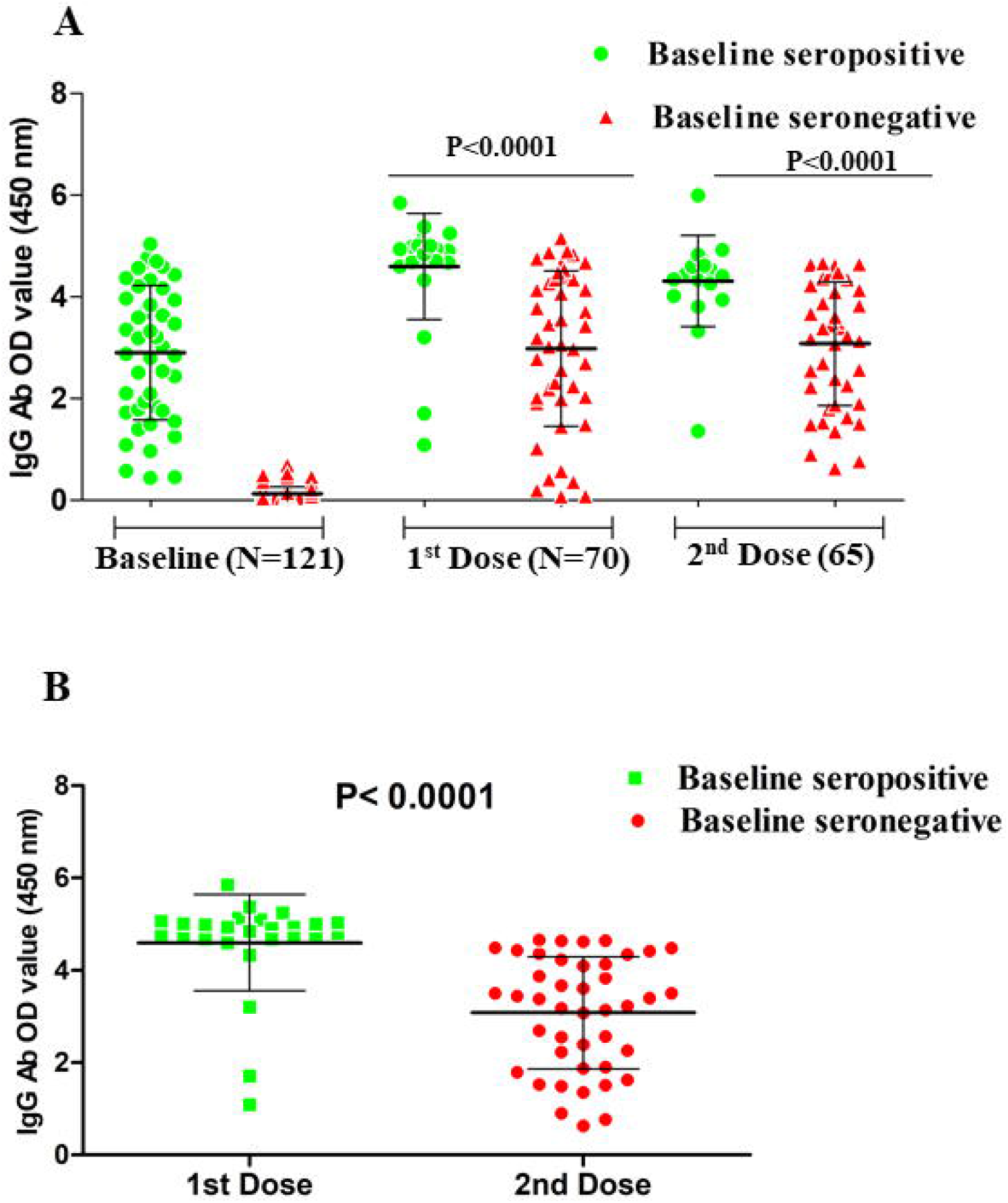
Represents scatter dot plot of IgG antibody titre against SARS-CoV-2 based on optical density measured at 450 nm among baseline seropositive and baseline seronegative subjects at three different time intervals of vaccination. Mean with ±SD is shown in each scatter dot plot. Figure 1A represents the comparison of IgG titre at baseline, after receiving of 1st and 2nd dose of vaccine for SARS-CoV2 among baseline seropositive and baseline seronegative study participants. Paired one tail students t-test was done for evaluation of IgG titre between study groups at three different time intervals. Figure 1B represents the IgG antibody titre among baseline seropositive subjects after receiving of 1st dose and baseline seronegative subjects after receiving of 2nd dose of vaccine. Unpaired students t-test revealed that IgG titres were significantly higher among baseline seropositive subjects after receiving 1st dose of vaccine than baseline seronegative subjects receiving the 2nd dose of vaccine.

Further studies on larger cohorts are needed to validate these findings, which are important for vaccine supply conservation and the introduction of systems to track vaccine response across a greater number of laboratories.

Therefore, for optimum vaccine usage and for developing ideal strategies to prevent further waves of the pandemic in India, vaccinees should be tested for SARS-CoV2 IgG antibodies before vaccination. As per a recent ICMR serosurvey study (3rd phase study pan-India with 28,589 samples screened) conducted in December 2020, the seroprevalence of SARS-CoV-2 in India was found to be 21.4% (unpublished data) [6]. This means that approximately 280 million people had been infected by December 2020 in India. Interestingly, the number of laboratory-confirmed cases in India by the end of 2020 was 10.03 million cases, which suggests that there are approximately 28 silent cases for every laboratory-confirmed case of COVID-19 in India. With this data, it can be extrapolated that by June 2, 2021, approximately 795 million populations (∼56.7%) of India’s population may already be infected with SARS-CoV-2. Therefore, SARS-CoV2 IgG antibody detection should be routinely done before vaccination. This will help in stratifying the previously infected population from the naive and can save up to 795 million doses of vaccines in India by introducing a single dose of vaccine for individuals with previous exposure and immunity against SARS-CoV2. Changing the present vaccine guideline to give only a single dose of Covishield vaccine to COVID-19 seropositive might help in meeting the increasing vaccine demand for vaccination. As a result, vaccination with both doses may be advised only for the vulnerable population or non-immune population.

## Conclusion

In participants with prior SARS-CoV2 infection, a single dose of ChAdOx1-nCOV (CovishieldTM®) vaccination may be adequate for a substantial immune response.

## Data Availability

The datasets generated for the current study are available with the corresponding author.

## Acknowledgement

The authors sincerely acknowledge all the study participants for consenting to the study. Authors also thank the Director, ICMR-Regional Medical Research Centre, NE Region, Assam, India.

## Declarations

### 3. Consent to participate

Written informed consent from each subject was obtained in before inclusion to the study.

### 4. Availability of data and material

The datasets generated for the current study are available with the corresponding author.

